# Identification and genomic analysis of pedigrees with exceptional longevity identifies candidate rare variants

**DOI:** 10.1101/2020.03.02.20030197

**Authors:** Justin B. Miller, Elizabeth Ward, Lyndsay A. Staley, Jeffrey Stevens, Craig C Teerlink, Justina P. Tavana, Matthew Cloward, Madeline Page, Louisa Dayton, Alzheimer’s Disease Genetics Consortium, Lisa A. Cannon-Albright, John S.K. Kauwe

**Author notes:** The authors wish it to be known that, in their opinion, the first two authors should be regarded as co-first authors. The authors wish it to be known that, in their opinion, the last two authors should be regarded as co-last authors.

## Abstract

**Background:** Longevity as a phenotype entails living longer than average and typically includes living without chronic age-related diseases. Recently, several common genetic components to longevity have been identified. This study aims to identify additional rare genetic variants associated with longevity using unique and powerful pedigree-based analyses of pedigrees with a statistical excess of healthy elderly individuals identified in the Utah Population Database (UPDB).

**Methods:** From an existing biorepository of Utah pedigrees, four pedigrees were identified which exhibited an excess of healthy elderly individuals; whole exome sequencing (WES) was performed on one set of elderly first- or second-cousins from each pedigree. Rare (<0.01 population frequency) variants shared by at least one elderly cousin pair in a region likely to be identical by descent were identified as candidates. Ingenuity Variant Analysis was used to prioritize putative causal variants based on quality control, frequency, and gain or loss of function. The variant frequency was compared in healthy cohorts and in an Alzheimer’s disease cohort. Remaining variants were filtered based on their presence in genes reported to have an effect on the aging process, aging of cells, or the longevity process. Validation of these candidate variants included tests of segregation to other elderly relatives.

**Results:** Fifteen rare candidate genetic variants spanning 17 genes shared within cousins were identified as having passed prioritization criteria. Of those variants, six were present in genes that are known or predicted to affect the aging process: *rs78408340* (*PAM*), *rs112892337* (*ZFAT*), *rs61737629* (*ESPL1*), *rs141903485* (*CEBPE*), *rs144369314* (*UTP4*), and *rs61753103* (*NUP88* and *RABEP1*). *ESPL1 rs61737629* and *CEBPE rs141903485* show additional evidence of segregation with longevity in expanded pedigree analyses (p-values=0.001 and 0.0001, respectively).

**Discussion:** This unique pedigree analysis efficiently identified several novel rare candidate variants that may affect the aging process and added support to seven genes that likely contribute to longevity. Further analyses showed evidence for segregation for two rare variants, *ESPL1 rs61737629* and *CEBPE rs141903485*, in the original longevity pedigrees in which they were originally observed. These candidate genes and variants warrant further investigation.

## INTRODUCTION

Aging is a major risk factor for various chronic diseases (Franceschi et al., 2018), but can also be considered as a phenotype (e.g. healthy aging with no chronic disease or exceptional longevity) (Lara et al., 2013). Genome-wide association studies have identified factors associated with longevity (Deelen et al., 2019; Pilling et al., 2017; Sebastiani et al., 2017). Genome-wide association studies identify associations between genotypes and phenotypes by testing individual genetic variants across a genome (Tam et al., 2019). However, they often lack sufficient power to identify rare variants because small effect sizes are diluted across thousands of individuals (Maher, 2008).

Pedigree-based analyses provide additional power to identify rare variants because they control for parent-of-origin effects, population stratification, and other hidden effects (Ott et al., 2011). Atzmon et al. (2006) capitalized on familial relationships in a case-control analysis of Ashkenazi Jews to identify variants specific to longevity. This study included 213 cases defined as individuals 95-107 years old living independently and in good health, and participants were required to have a child participate in the study. The offspring group consisted of 216 individuals and a control group consisted of 258 individuals. This study suggested that pathways involved in lipoprotein metabolism appear to influence longevity in humans.

An additional study on longevity was conducted as part of the Hawaii Lifespan Study, and included healthy elderly individuals from the original population of the Honolulu Heart Program and Honolulu Asia Aging Study (Willcox et al., 2008). The Honolulu Heart Program is a population-based, prospective study that began in 1965 by studying cardiovascular disease among 8,006 Japanese American men. This study contained 213 cases who survived to at least 95 years of age. The mean age of death for the 402 control individuals in the Honolulu Asia Aging Study and the Hawaii Lifespan Study who died near the mean death age for the 1910 U.S. birth cohort was 78.5 years of age. This study identified common, natural genetic variation strongly associated with longevity in the *FOXO3A* gene.

The Long Life Family study also contains a multi-center family-based cohort that was used to identify genetic components of longevity. This study demonstrated the use of sequencing within pedigrees to identify 24 inherited rare variants in two long-lived families influencing healthy aging (Druley et al., 2016).

The Utah Population Database (UPDB) includes extensive sets of demographic and medical records for more than 11 million individuals, three million of whom are linked to Utah pedigree data (Cannon Albright, 2008). From an existing biorepository of stored DNA for Utah individuals identified in the UPDB spanning decades, clusters of related sampled healthy elderly individuals (age at death greater than 90 years) were identified. Sampled elderly cousin pairs selected from four of these pedigrees, which exhibited a statistical excess of individuals who died at an age older than 90 years, were sequenced. Putative causal variants were identified using an efficient and powerful analytical approach previously used to identify rare variants that influence risk and resilience to Alzheimer’s disease (Patel et al., 2019; Ridge et al., 2017), melanoma (Teerlink et al., 2018), Osteoporosis (Teerlink et al., 2020), and colorectal cancer (Thompson et al., 2020) in UPDB pedigrees.

## MATERIALS AND METHODS

### Data

#### Utah Population Database (UPDB)

The UPDB includes population-based resources that link computerized demographic and health data with the electronic genealogical records of the 18^th^ century founders of Utah and their descendants to modern day (Cannon Albright, 2008). The multigenerational pedigrees represented in UPDB were constructed from data provided by the Genealogical Society of Utah and have been expanded extensively based on Utah State vital records. There are currently over 11 million individuals included in the database, including approximately three million people with at least three generations of family history connected to the original Utah settlers. Age at death was calculated from death dates provided in genealogy records and from over 900,000 death certificates linked to the UPDB genealogy.

#### Longevity Pedigrees

Analyses were performed on approximately 36,000 individuals from the UPDB for whom DNA is available and compared against high-risk cancer pedigrees. All clusters of related sampled healthy elderly individuals (age at death greater than 90 years; consented and sampled for research at age greater than 85 years) were identified. Four of these sampled pedigrees with a statistical excess of individuals who died at an age older than 90 years that also included at least one sampled healthy elderly cousin pair were selected for analysis. One such individual was a member of two independent pedigrees, through different ancestors, and one pedigree included three related sampled cousins for a total of eight individuals analyzed.

#### Alzheimer’s Disease Genetic Consortium

Various analyses were conducted using the Alzheimer’s Disease Genetic Consortium (ADGC) datasets compiled by Naj et al. (2011). ADGC is a collection of 30 merged datasets spanning 1984 to 2012, and was established to help identify genetic markers of late-onset Alzheimer’s disease (Boehme et al., September 2014). ADGC contains imputed SNP array data for 28,730 subjects (58.34% female), including 10,486 Alzheimer’s disease cases and 10,168 healthy controls. ADGC imputed the 30 datasets to the Haplotype Reference Consortium (HRC) reference panel, which includes 64,976 haplotypes and 39,235,157 SNPs (Loh et al., 2016; Naj et al., 2017). Genotyped markers with a minor allele frequency less than 0.01 and markers that deviated from Hardy Weinberg Equilibrium were removed. All aspects of the study were approved by institutional review boards, and each applicant signed a written form of consent for their genetic data to be used for research purposes.

#### The Wellderly Study

The Wellderly Study is a cohort of more than 1,400 individuals over the age of 80 with no chronic disease or chronic use of medication (Erikson et al., 2016). The purpose of this study was to determine whether genetic factors underlie the phenotype of exceptional longevity. Researchers performed whole genome sequencing on 511 Wellderly participants and compared their results to whole genome sequencing data from 686 young adults from the Inova Translational Medicine Institute (ITMI), which served as an ethnicity-matched population control (Bodian et al., 2014).

Wellderly individuals had significantly reduced genetic risk for coronary artery disease (p-value=2.54 × 10^−3^) and Alzheimer’s disease (p-value=9.84 × 10^−4^), and no decrease in the rate of rare pathogenic variants. These findings suggest the presence of other disease-resistant factors within this longevity cohort.

#### Bioinformatic Analysis

Whole exome sequencing for the eight elderly individuals selected as cousin pairs was performed at the Huntsman Cancer Institute’s Genomics Core facility. A DNA library was prepared from 2µg of DNA per sample using the Agilent SureSelect XT Human All Exon + UTR (v5) capture kit. Samples were run on the Illumina HiSeq 2000 sequencer that generates paired-end reads of up to 150 base pairs in length. Raw reads were mapped to the human genome v37 (GRCh37) reference genome using BWA-MEM (Li, 2013; Li and Durbin, 2009). Variants were called using Genome Analysis Toolkit 3.5.0 (GATK) (McKenna et al., 2010) software following Broad Institute Best Practices Guidelines. Variants occurring outside the exon capture kit intended area of coverage were removed. Variants were annotated with ANNOVAR (Wang et al., 2010). Candidate variants were filtered on the criteria of being rare in population (minor allele frequency less than 0.01) and shared by a cousin pair.

#### Genetic Support for Pedigree Enrichment

In order to evaluate the effectiveness of pedigree enrichment for longevity, a polygenic risk score analysis was conducted for each of the eight individuals in the dataset. A polygenic risk score calculates the cumulative risk for a certain phenotype determined from aggregating the effect sizes of multiple genetic loci (Sugrue and Desikan, 2019). The polygenic risk score was calculated from the following equation, where *a*_*i*_ is the number of alleles at the *i*^*th*^ locus, *r*_*i*_ is the odds ratio at the *i*^*th*^ locus, and *p* is the p-value of the odds ratio:

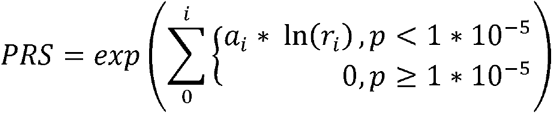

For each sample, the polygenic risk score for Alzheimer’s disease was calculated using the odds ratios from Lambert et al. (2013), coronary artery disease using the odds ratios from Schunkert et al. (2011), and heart failure using the odds ratios from Shah et al. (2020). The same genome-wide association studies were used to calculate polygenic risk scores for each individual in the ADGC controls.

#### Segregation Validation using Rare Variant Sharing

Candidate variants were assayed with TaqMan in a set of 120 sampled individuals who died after 90 years of age. These individuals were members of the four extended longevity pedigrees from which the original elderly cousins were identified and were included in 180 additional sampled individuals who died after 95 years of age. The *RVsharing* program (Bureau et al., 2014) was used to statistically assess segregation of candidate rare variants in other sampled affected relatives. *RVsharing* calculates the probability of seeing rare variants in the observed pattern of carriage for a specified pedigree structure based on a relatedness matrix between cases, based on genealogy data. A p-value threshold of 0.05 effectively discriminates between rare variants that segregate (Teerlink et al., 2016).

## RESULTS

Whole exome sequencing data was generated for elderly cousin pairs in four pedigrees with a statistical excess of long-lived individuals. Using UPDB pedigrees to identify candidate predisposition variants for a phenotype of interest allows efficient generation of the set of rare variants that are shared in related (typically cousin) pairs of individuals with the phenotype of interest who are also members of pedigrees that have been established to be at “high-risk” for the phenotype. Since the affected cousin pairs are members of the same high-risk pedigree, they are hypothesized to share the predisposition variant of interest. The set of rare variants shared in any of the cousin pairs from the high-risk pedigrees therefore constitute likely candidate predisposition variants. Using a small set of four “high-risk longevity” pedigrees, 83 rare variants with a minor allele frequency less than 0.01 in the general population that were shared within at least one cousin pair were efficiently identified.

### Polygenic Risk Score Analysis

Figure 2 displays the distribution of polygenic risk scores for Alzheimer’s disease, coronary artery disease, and heart failure in the longevity dataset (n=8) against the distribution of risk scores for ADGC controls (n=13,410).Although the cousins are related, they share a relatively low proportion of their genomes (12.5% for first cousins and 3.13% for second cousins), which allows most common variants used in calculating polygenic risk scores to maintain the same degree of independence between cousins as between unrelated individuals. In all but one instance, the most similar polygenic risk score for an individual in the dataset for any of the three tested diseases was not with their cousin, butwith a different unrelated individual in the dataset. Therefore, a Welch’s two-sample t-test was performed to reveal a significant difference between the mean scores of the longevity cousin pairs and the ADGC controls for coronary artery disease (t=-30.192; p-value = 7.35×10^−9^) and heart failure (t=-21.746; p-value = 9.78 × 10^−8^). These analyses indicate that the cousin pairs have fewer common variants that contribute to common diseases in elderly individuals than the ADGC control group,suggesting that the pedigree identification effectively selected families enriched with exceptional longevity due to decreased risk for disease. Supplemental Table S1 outlines the polygenic risk scores for each individual in the dataset, including the prioritized variants present in each person.

**Figure 1:**
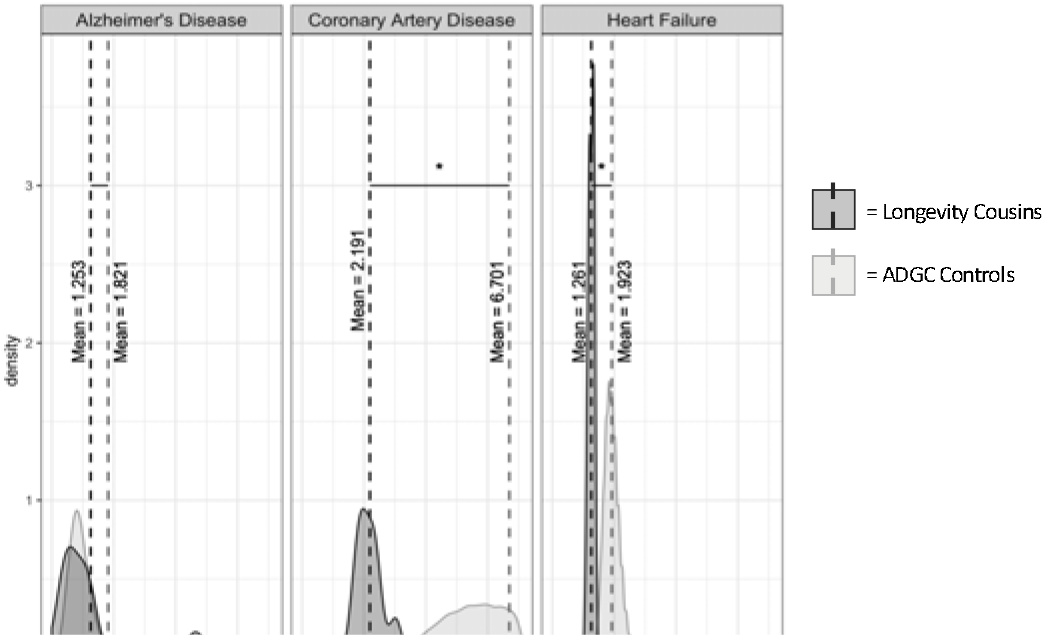
Polygenic Risk Scores for UPDB cousins. The distribution of risk scores for the longevity cousins are plotted against the polygenic risk score distribution of the ADGC controls.

**Figure 2:**
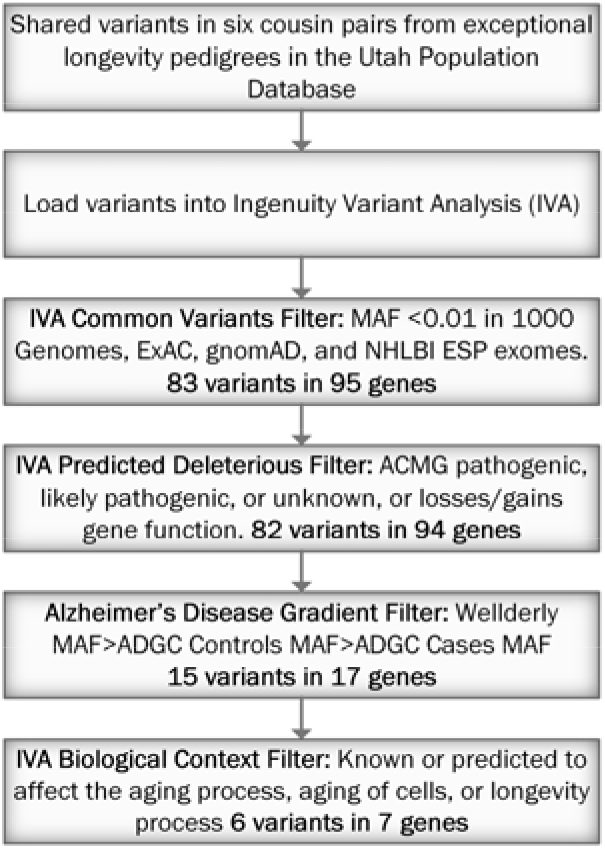
Pipeline for Rare Variant Analysis in Cousin Pairs. Flowchart explaining the filters that we used on our dataset, including the number of variants and genes that passed each filter.

#### Variant Prioritization

A rare variant analysis was performed on the cousin pairs by first limiting selection to variants that were shared by at least one cousin pair. A Common Variants Filter in Ingenuity® Variant Analysis™ software from QIAGEN, Inc. was used to remove all variants with a minor allele frequency greater than 0.01 in 1000 Genomes (Auton et al., 2015), Exome Aggregation Consortium (ExAC) (Karczewski et al., 2017), The Genome Aggregation Database (gnomAD) (Karczewski et al., 2019), or the NHLBI GO Exome Sequencing Project (ESP), Seattle, WA (URL: http://evs.gs.washington.edu/EVS/) [March 2018]. This step identified 83 rare candidate variants spanning 95 genes, including 12 variants that each affect two genes. A series of filtration methods on these 83 variants using Ingenuity Variant Analysis was used to prioritize a candidate list of variants associated with longevity (see Figure 1). Variants remaining after each filter are listed in Supplementary File S1.

#### Predicted Deleterious Filter

After the Common Variants Filter, the Predicted Deleterious Filter in Ingenuity Variant Analysis was applied to select variants that were associated with the loss or gain of gene function or were considered ‘Pathogenic’, ‘Likely Pathogenic’, or ‘Unknown’ according to the American College of Medical Genetics and Genomics (ACMG) Guidelines for variant classification (Richards et al., 2015). This analysis excluded only one variant, refining the list to 82 variants spanning 94 genes.

#### Alzheimer’s Disease Risk Gradient Filter

The purpose of this filter was to identify rare variants that are present more frequently in healthy cohorts than diseased cohorts, since it is expected that protective rare variants that positively impact longevity will not be present as frequently in diseased cohorts. Each variant was compared to the Wellderly dataset and the ADGC dataset to ensure that variants followed expected population allele frequencies based on the number of healthy individuals in each elderly cohort. For this filter, the minor allele frequency of each rare variant was required to be higher in the Wellderly cohort than the ADGC control group, and have a higher minor allele frequency higher in the ADGC control group than the ADGC Alzheimer’s disease cases. Genetic variants that passed this filter indicated a higher variant occurrence in healthy individuals than diseased individuals. Fifteen variants spanning 17 genes passed this filter.

#### Biological Context Filter

The final filter evaluated the biological function of each of the 15 remaining variants. This filter included only variants in genes that were known or predicted to affect the aging process, aging of cells, or the longevity process. This filter prioritized six variants spanning seven genes.Recognizing that the biological context filter depends on an accurate understanding of the biological functions of each of the 17 genes that passed the Alzheimer’s Disease Risk Gradient Filter, it is possible that all 15 candidate variants that passed the Alzheimer’s Disease Risk Gradient Filter also positively affect longevity. However, the following six variants that passed the Biological Context Filter are the most supported candidate variants: *rs78408340* (*PAM*), *rs112892337* (*ZFAT*), *rs61737629* (*ESPL1*), *rs141903485* (*CEBPE*), *rs144369314* (*UTP4*), and *rs61753103* (*NUP88* and *RABEP1*).

#### Rare Variant Segregation Analysis

Two rare variants passing all filters were also pursued with segregation analysis. *ESPL1 rs61737629* was selected because it was the only variant to be observed in more than one cousin pair, and *CEBPE rs141903485* was selected because it has a regulomeDB score of 2b. These two variants were assayed in 213 additional healthy elderly individuals (sampled after age 90 years) and 182 sampled Alzheimer’s disease cases (confirmed by Utah death certificate) from the UPDB. The two variants were also assayed in 11 additional longevity samples in the pedigree in which both of the variants were originally observed. *ESPL1 rs61737629* was observed in four additional longevity cases. *CEBPE rs141903485* was observed in seven additional longevity cases and three Alzheimer’s disease cases. Additional analyses in the original longevity pedigree in which both variants were identified also identified one more carrier of *ESPL1 rs61737629* and three additional carriers of *CEBPE rs141903485*. The Rare Variant Sharing test for *ESPL1 rs61737629* (p-value = 0.001) and *CEBPE rs141903485* (p-value = 0.0001) reveal that there is a low probability of these variants being shared within healthy elderly individuals in this pedigree by random chance. The constellation of variant carriers of *ESPL1 rs61737629* and *CEBPE rs141903485* within this extended pedigree was used to calculate the Rare Variant Sharing value for each variant and provides statistical evidence that *ESPL1 rs61737629* and *CEBPE rs141903485* segregate significantly with longevity.

## DISCUSSION

### Prioritized Variants

Familial relationships and previously sampled individuals ascertained in the UPDB were leveraged to identify rare candidate variants that influence exceptional longevity. The rare variant analysis pipeline identified six candidate variants located in seven genes that demonstrate a convincing case for association with longevity (see Table 1).

**Table 1:** Final Six Prioritized Variants associated with Longevity from the Six Cousin Pairs. This table shows the results of the final Ingenuity Variant Analysis Biological Context Filter.

| Chromosome | Position in GRCh37 | Reference | Alternate | Accession Number | Gene Name | SIFT Function Prediction | Translation Impact |
| --- | --- | --- | --- | --- | --- | --- | --- |
| 5 | 102338739 | C | G | *rs78408340* | *PAM* | Damaging | missense |
| 8 | 135614553 | G | C | *rs112892337* | *ZFAT* | Damaging | missense |
| 12 | 53682043 | C | G | *rs61737629* | *ESPL1* | Damaging | missense |
| 14 | 23587838 | G | T | *rs141903485* | *CEBPE* | Damaging | missense |
| **16** | 69170741 | G | T | *rs144369314* | *UTP4* | Tolerated | missense |
| **17** | 5289554 | T | C | *rs61753103* | *NUP88, RABEP1* | Tolerated | missense |

Missense mutation *rs78408340* in the *PAM* gene was identified to have potential association with longevity and is categorized by SIFT (Sim et al., 2012) as ‘Damaging.’ *PAM* catalyzes the conversion of neuroendocrine peptides to active alpha-amidated products. Alleles associated with type-2 diabetes in *PAM*, including *rs78408340*, reduce the gene’s function, which alters the amidation of peptides critical for insulin secretion. Therefore, *rs78408340*, along with other alleles in *PAM*, confers higher risk for type-2 diabetes (Fuchsberger et al., 2016; Steinthorsdottir et al., 2014). One cousin pair shared the variant *PAM rs78408340*, which may account for these individuals’ shared phenotype.

The individuals in the same cousin pair are also carriers of the variant *rs112892337* in the *ZFAT* gene, which is also labelled by SIFT as ‘Damaging.’ Little is known about the function of this specific allele. However, *ZFAT* is expressed in B and T lymphocytes and has shown to be a critical transcription regulator involved in apoptosis and cell survival (Fujimoto et al., 2009).Bourguiba-Hachemi et al. (2016) found that another variant, *rs733254*, in *ZFAT* is a risk marker for multiple sclerosis (MS) in women. Multiple studies have also detected an association between *ZFAT* and the severity of autoimmune thyroid disease (Inoue et al., 2012; Sakai et al., 2001).

Missense mutation *rs61737629* in *ESPL1* was prioritized by the filtration pipeline and shared by two cousin pairs. SIFT also predicts this variant to be ‘Damaging.’ *ESPL1*, which encodes separase, initiates the final separation of sister chromatids before anaphase by cleaving the subunit SCC1. Disruption of the separase function leads to chromosomal instability, and abnormal expression of this gene results in severe medical consequences. Due to the overexpression of separase in luminal tumors, *ESPL1* is a promising candidate oncogene in luminal cancers (Finetti et al., 2014). Currently, the behavior of *ESPL1 rs61737629* is unknown. This study may lend additional support to luminal cancer studies exploring this variant.

Three individuals, representing two independent cousin pairs, shared *CEBPE rs141903485*, a missense variant labelled as ‘Damaging’ by SIFT. *CEBPE* encodes a bZIP transcription factor and plays a role in gene regulation in myeloid and lymphoid lineages (Antonson et al., 1996). The loss of *CEBPE* function influences the pathogenesis of myeloid disorders, including acute myeloid leukemia (Truong et al., 2003) and pediatric B-cell acute lymphoblastic leukemia (Gharbi et al., 2016; Studd et al., 2019; Sun et al., 2015; Wang et al., 2015). The variant *rs141903485* is associated with pediatric B-cell acute lymphoblastic leukemia susceptibility (Xu et al., 2013; Xu et al., 2015).

The missense variant *rs144369314* located in *UTP4* was shared by one cousin pair. *UTP4* encodes a WD40-repeat-containing protein that is localized to the nucleolus. Variation in *UTP4* is significantly associated with North American Indian childhood cirrhosis (Freed and Baserga, 2010; Yu et al., 2005).

Individuals in one cousin pair carry the missense mutation *rs61753103* implicated in the gene *NUP88. NUP88* regulates the flow of macromolecules between the nucleus and the cytoplasm, is overexpressed in malignancies, and is considered a putative marker for tumor growth (Hashizume et al., 2010; Lang et al., 2017; Martinez et al., 1999). Increased expression of this gene is associated with tumor aggressiveness in uterine and breast cancer (Agudo et al., 2004; Schneider et al., 2010) and higher risk for colorectal cancer (Zhao et al., 2012).

This same variant, *rs61753103*, is located in the *RABEP1* gene. *RABEP1* is involved in endocytic membrane fusion and membrane trafficking. A recent genome-wide association study identified *RABEP1* to be associated with increased Alzheimer’s disease risk (Jansen et al., 2018).

Most of the prioritized variants identified here are located in genes that directly affect chronic diseases. While additional biological validation is required to better characterize the relationship between these loci and the longevity process, it is promising that the prioritized variants are located on genes previously implicated in disease.

### Variants in Previously Identified Longevity Candidate Genes

Strict filters were used to identify the most likely causal variants in this set of four longevity pedigrees. However, the filtering criteria likely contribute to a high false negative rate and therefore it is unlikely that this analysis has provided an exhaustive list of all variants associated with longevity in these pedigrees. Furthermore, the use of whole exome sequencing data limits the ability to detect any significant variants that reside outside the protein-coding regions of genes. Five additional variants that were shared in at least one cousin pair were identified in genes previously implicated in longevity: *PROX2, SEMA6D, MARK4, MEF2A*, and *EBF1*.*PROX2* is a transcription factor specific to RNA polymerase II implicated in lens fiber cell morphogenesis and lymphatic endothelial cell differentiation and associated with parental longevity (Pilling et al., 2017). One cousin pair carried a frameshift variant at position 75321938 on chromosome 14 (no accession) implicated in this locus. This variant was not prioritized here because there was information about its frequency in the ADGC dataset.

Pilling et al. (2017) also identified variation in *SEMA6D* associated with longer parental lifespan. *SEMA6D* is involved in the immune response, and is responsible for the maintenance and modification of neuronal connections (He et al., 2002). Multiple studies have found *SEMA6D* to be related to tumor angiogenesis and to play an important role in the development of gastric cancer (Qu et al., 2019; Zhao et al., 2006). One cousin pair shared the missense mutation *rs769450413* located in this gene. However, the Alzheimer’s Disease Risk Gradient Filter also failed to prioritize this variant because it was not genotyped in the ADGC dataset.

*MARK4* regulates the transition between stable and dynamic microtubules and plays a role in cell cycle progression (Rovina et al., 2014). *MARK4* also regulates tau protein phosphorylation and is proposed to be functionally important to the progression of Alzheimer’s disease (Gu et al., 2013; Seshadri et al., 2010; Sun et al., 2016) and parental longevity (Pilling et al., 2017). Multiple studies also provide evidence for the expression of *MARK4* as a potential marker for breast and prostate cancer (Heidary Arash et al., 2017; Jenardhanan et al., 2014; Pardo et al., 2016). One cousin pair shared the missense variant *rs753496642* in this gene, which SIFT categorizes as ‘Damaging.’ This mutation was also excluded by the Alzheimer’s Disease Risk Gradient Filter because there was no information about its frequency in the ADGC dataset.

*MEF2A* conveys significant association with healthy aging (Druley et al., 2016). *MEF2A* is a transcriptional activator involved in muscle development, neuronal differentiation, cell growth control, and apoptosis. Variants in the 3’-UTR region of this gene are associated with coronary artery disease (Huang and Wang, 2015; Xiong et al., 2019; Xu et al., 2016). *EBF1* is a transcriptional activator which identifies changes in the palindromic sequence. *EBF1* is involved in the regulation of metabolic and inflammatory signaling pathways, and the loss of gene function results in impaired insulin and inflammatory signaling (Griffin et al., 2013). *EBF1* plays a role in a variety of diseases including breast cancer (Fernandez-Jimenez et al., 2017; Garcia-Closas et al., 2013; Michailidou et al., 2013), coronary artery disease (Ehret et al., 2011; Li et al., 2017; Singh et al., 2015; Wain et al., 2011), Hodgkin lymphoma (Bohle et al., 2013), multiple sclerosis (Martinez et al., 2005; Sombekke et al., 2010), and leukemia (Heltemes-Harris et al., 2011; Mesuraca et al., 2015; Welsh et al., 2018). *MEF2A* and *EBF1* are regulators for the *DMAC2* gene, which was implicated in one cousin pair. The *DMAC2* variant, *rs139204637*, passed all but the Biological Context filter, because *DMAC2* has not previously been implicated in the aging process.

Efforts to understand the genetic basis of longevity phenotypes have yielded few definitive findings to date. As is the case with other traits, heterogeneity in the diagnosis and etiology of these phenotypes creates significant challenges. For example, longevity is clearly influenced by genetics, epigenetics, environment, and chance (e.g., no fatal accidents early in life). The high-risk pedigree-based approach minimizes genetic heterogeneity and may also reduce other sources of heterogeneity; recall bias was reduced by the existence of extensive genealogy data. This analysis of whole exome sequences in longevity pedigrees identified six putative causal variants,including two that showed evidence of segregation in extended pedigree analyses. Biological validation of these candidates is necessary to characterize variant effects, the filtering criteria used might have allowed for false positive results due to chance sharing of rare variants among relatives. These findings suggest that further evaluation of these candidate variants is warranted and highlight the utility of this unique pedigree-based approach to gene discovery.

## Data Availability

Data from the UPDB, the ADGC, and the Wellderly study were obtained using the applicable request forms and handled in accordance with IRB-approved methods.

## Acknowledgements

We appreciate the contributions of Brigham Young University in supporting this research. This research is supported by RF1AG054052 (PI: Kauwe) and U01AG052411 (PI: Goate).

We thank the Pedigree and Population Resource of Huntsman Cancer Institute, University of Utah (funded in part by the Huntsman Cancer Foundation) for its role in the ongoing collection, maintenance and support of the Utah Population Database (UPDB). We also acknowledge partial support for the UPDB through grant P30 CA2014 from the National Cancer Institute, University of Utah and from the University of Utah’s program in Personalized Health and Center for Clinical and Translational Science.

The authors would like to thank the NHLBI GO Exome Sequencing Project and its ongoing studies which produced and provided exome variant calls for comparison: the Lung GO Sequencing Project (HL-102923), the WHI Sequencing Project (HL-102924), the Broad GO Sequencing Project (HL-102925), the Seattle GO Sequencing Project (HL-102926) and the Heart GO Sequencing Project (HL-103010).

Alzheimer’s Disease Genetics Consortium (ADGC)

Data from ADGC was appropriately downloaded from dbGaP (accession: phs000372.v1.p1). We acknowledge the contributions of

The members of the Alzheimer’s Disease Genetics Consortium are: Marilyn S. Albert^1^, Roger L. Albin^2-4^, Liana G. Apostolova^5^, Steven E. Arnold^6^, Clinton T. Baldwin^7^, Robert Barber^8^, Michael M. Barmada^9^, Lisa L. Barnes^10, 11^, Thomas G. Beach^12^, Gary W. Beecham^13, 14^, Duane Beekly^15^, David A. Bennett^10, 16^, Eileen H. Bigio^17^, Thomas D. Bird^18^, Deborah Blacker^19,20^, Bradley F. Boeve^21^, James D. Bowen^22^, Adam Boxer^23^, James R. Burke^24^, Joseph D. Buxbaum^25, 26, 27^, Nigel J. Cairns^28^, Laura B. Cantwell^29^, Chuanhai Cao^30^, Chris S. Carlson^31^, Regina M. Carney^13^, Minerva M. Carrasquillo^33^, Steven L. Carroll^34^, Helena C. Chui^35^, David G. Clark^36^, Jason Corneveaux^37^, Paul K. Crane^38^, David H. Cribbs^39^, Elizabeth A. Crocco^40^, Carlos Cruchaga^41^, Philip L. De Jager^42,43^, Charles DeCarli^44^, Steven T. DeKosky^45^, F. Yesim Demirci^9^, Malcolm Dick^46^, Dennis W. Dickson^33^, Ranjan Duara^47^, Nilufer Ertekin-Taner^33,48^, Denis Evans^49^, Kelley M. Faber^50^, Kenneth B. Fallon^34^, Martin R. Farlow^51^, Lindsay A Farrer^7,52,76,77,83^, Steven Ferris^53^, Tatiana M. Foroud^50^, Matthew P. Frosch^54^, Douglas R. Galasko^55^, Mary Ganguli^56^, Marla Gearing^57,58^, Daniel H. Geschwind^59^, Bernardino Ghetti^60^, John R. Gilbert^13,14^, Sid Gilman^2^, Jonathan D. Glass^61^, Alison M. Goate^41^, Neill R. Graff-Radford^33,48^, Robert C. Green^62^, John H. Growdon^63^, Jonathan L. Haines^64, 65^, Hakon Hakonarson^66^, Kara L. Hamilton-Nelson^13^, Ronald L. Hamilton^67^, John Hardy^68^, Lindy E. Harrell^36^, Elizabeth Head^69^, Lawrence S. Honig^70^, Matthew J. Huentelman^37^, Christine M. Hulette^71^, Bradley T. Hyman^63^, Gail P. Jarvik^72,73^, Gregory A. Jicha^74^, Lee-Way Jin^75^, Gyungah Jun^7,76,77^, M. Ilyas Kamboh^9,78^, Anna Karydas^23^, John S.K. Kauwe^79^, Jeffrey A. Kaye^80,81^, Ronald Kim^82^, Edward H. Koo^55^, Neil W. Kowall^83,84^, Joel H. Kramer^85^, Patricia Kramer^80,86^, Walter A. Kukull^87^, Frank M. LaFerla^88^, James J. Lah^61^, Eric B. Larson^38,89^, James B. Leverenz^90^, Allan I. Levey^61^, Ge Li^91^, Andrew P. Lieberman^92^, Chiao-Feng Lin^29^, Oscar L. Lopez^78^, Kathryn L. Lunetta^76^, Constantine G. Lyketsos^93^, Wendy J. Mack^94^, Daniel C. Marson^36^, Eden R. Martin^13,14^, Frank Martiniuk^95^, Deborah C. Mash^96^, Eliezer Masliah^55,97^, Richard Mayeux^70, 109, 110^, Wayne C. McCormick^38^, Susan M. McCurry^98^, Andrew N. McDavid^31^, Ann C. McKee^83,84^, Marsel Mesulam^99^, Bruce L. Miller^23^, Carol A. Miller^100^, Joshua W. Miller^75^, Thomas J. Montine^90^, John C. Morris^28, 101^, Jill R. Murrell^50, 60^, Amanda J. Myers^40^, Adam C. Naj^13^, John M. Olichney^44^, Vernon S. Pankratz^102^, Joseph E. Parisi^103,104^, Margaret A. Pericak-Vance^13, 14^, Elaine Peskind^91^, Ronald C. Petersen^21^, Aimee Pierce^39^, Wayne W. Poon^46^, Huntington Potter^30^, Joseph F. Quinn^80^, Ashok Raj^30^, Murray Raskind^91^, Eric M. Reiman^37,105-107^, Barry Reisberg^53,108^, Christiane Reitz^70,109,110^, John M. Ringman^5^, Erik D. Roberson^36^, Ekaterina Rogaeva^111^, Howard J. Rosen^23^, Roger N. Rosenberg^112^, Mary Sano^26^, Andrew J. Saykin^50,113^, Gerard D. Schellenberg^29^, Julie A. Schneider^10,114^, Lon S. Schneider^35,115^, William W. Seeley^23^, Amanda G. Smith^30^, Joshua A. Sonnen^90^, Salvatore Spina^60^, Peter St George-Hyslop^111,116^, Robert A. Stern^83^, Rudolph E. Tanzi^63^, John Q. Trojanowski^29^, Juan C. Troncoso^117^, Debby W. Tsuang^91^, Otto Valladares^29^, Vivianna M. Van Deerlin^29^, Linda J. Van Eldik^118^, Badri N. Vardarajan^7^, Harry V. Vinters^5,119^, Jean Paul Vonsattel1^20^, Li-San Wang^29^, Sandra Weintraub^99^, Kathleen A. Welsh-Bohmer^24, 121^, Jennifer Williamson^70^, Randall L. Woltjer^122^, Clinton B. Wright^123^, Steven G. Younkin^33^, Chang-En Yu^38^, Lei Yu^10^

^1^Department of Neurology, Johns Hopkins University, Baltimore, Maryland, ^2^Department of Neurology, University of Michigan, Ann Arbor, Michigan, ^3^Geriatric Research, Education and Clinical Center (GRECC), VA Ann Arbor Healthcare System (VAAAHS), Ann Arbor, Michigan, ^4^Michigan Alzheimer Disease Center, Ann Arbor, Michigan, ^5^Department of Neurology, University of California Los Angeles, Los Angeles, California, ^6^Department of Psychiatry, University of Pennsylvania Perelman School of Medicine, Philadelphia, Pennsylvania, ^7^Department of Medicine (Genetics Program), Boston University, Boston, Massachusetts, ^8^Department of Pharmacology and Neuroscience, University of North Texas Health Science Center, Fort Worth, Texas, ^9^Department of Human Genetics, University of Pittsburgh, Pittsburgh, Pennsylvania, ^10^Department of Neurological Sciences, Rush University Medical Center, Chicago, Illinois, ^11^Department of Behavioral Sciences, Rush University Medical Center, Chicago, Illinois, ^12^Civin Laboratory for Neuropathology, Banner Sun Health Research Institute, Phoenix, Arizona, ^13^The John P. Hussman Institute for Human Genomics, University of Miami, Miami, Florida, ^14^Dr. John T. Macdonald Foundation Department of Human Genetics, University of Miami, Miami, Florida, ^15^National Alzheimer’s Coordinating Center, University of Washington, Seattle, Washington, ^16^Rush Alzheimer’s Disease Center, Rush University Medical Center, Chicago, Illinois, ^17^Department of Pathology, Northwestern University, Chicago, Illinois, ^18^Department of Neurology, University of Washington, Seattle, Washington, ^19^Department of Epidemiology, Harvard School of Public Health, Boston, Massachusetts, ^20^Department of Psychiatry, Massachusetts General Hospital/Harvard Medical School, Boston, Massachusetts, ^21^Department of Neurology, Mayo Clinic, Rochester, Minnesota, ^22^Swedish Medical Center, Seattle, Washington, ^23^Department of Neurology, University of California San Francisco, San Francisco, California, ^24^Department of Medicine, Duke University, Durham, North Carolina, ^25^Department of Neuroscience, Mount Sinai School of Medicine, New York, New York, ^26^Department of Psychiatry, Mount Sinai School of Medicine, New York, New York, ^27^Departments of Genetics and Genomic Sciences, Mount Sinai School of Medicine, New York, New York, ^28^Department of Pathology and Immunology, Washington University, St. Louis, Missouri, ^29^Department of Pathology and Laboratory Medicine, University of Pennsylvania Perelman School of Medicine, Philadelphia, Pennsylvania, ^30^USF Health Byrd Alzheimer’s Institute, University of South Florida, Tampa, Florida, ^31^Fred Hutchinson Cancer Research Center, Seattle, Washington, ^32^Department of Psychiatry, Vanderbilt University, Nashville, Tennessee, ^33^Department of Neuroscience, Mayo Clinic, Jacksonville, Florida, ^34^Department of Pathology, University of Alabama at Birmingham, Birmingham, Alabama, ^35^Department of Neurology, University of Southern California, Los Angeles, California, ^36^Department of Neurology, University of Alabama at Birmingham, Birmingham, Alabama, ^37^Neurogenomics Division, Translational Genomics Research Institute, Phoenix, Arizona, ^38^Department of Medicine, University of Washington, Seattle, Washington, 39Department of Neurology, University of California Irvine, Irvine, California, ^40^Department of Psychiatry and Behavioral Sciences, Miller School of Medicine, University of Miami, Miami, Florida, ^41^Department of Psychiatry and Hope Center Program on Protein Aggregation and Neurodegeneration, Washington University School of Medicine, St. Louis, Missouri, ^42^Program in Translational NeuroPsychiatric Genomics, Institute for the Neurosciences, Department of Neurology & Psychiatry, Brigham and Women’s Hospital and Harvard Medical School, Boston, Massachusetts, ^43^Program in Medical and Population Genetics, Broad Institute, Cambridge, Massachusetts, ^44^Department of Neurology, University of California Davis, Sacramento, California, 45University of Virginia School of Medicine, Charlottesville, Virginia, ^46^Institute for Memory Impairments and Neurological Disorders, University of California Irvine, Irvine, California, ^47^Wien Center for Alzheimer’s Disease and Memory Disorders, Mount Sinai Medical Center, Miami Beach, Florida, ^48^Department of Neurology, Mayo Clinic, Jacksonville, Florida, ^49^Rush Institute for Healthy Aging, Department of Internal Medicine, Rush University Medical Center, Chicago, Illinois, ^50^Department of Medical and Molecular Genetics, Indiana University, Indianapolis, Indiana, ^51^Department of Neurology, Indiana University, Indianapolis, Indiana, ^52^Department of Epidemiology, Boston University, Boston,Massachusetts, ^53^Department of Psychiatry, New York University, New York, New York, ^54^C.S. Kubik Laboratory for Neuropathology, Massachusetts General Hospital, Charlestown, Massachusetts, ^55^Department of Neurosciences, University of California San Diego, La Jolla, California, ^56^Department of Psychiatry, University of Pittsburgh, Pittsburgh,Pennsylvania, ^57^Department of Pathology and Laboratory Medicine, Emory University, Atlanta, Georgia, ^58^Emory Alzheimer’s Disease Center, Emory University, Atlanta, Georgia, ^59^Neurogenetics Program, University of California Los Angeles, Los Angeles, California, ^60^Department of Pathology and Laboratory Medicine, Indiana University, Indianapolis, Indiana, ^61^Department of Neurology, Emory University, Atlanta, Georgia, ^62^Division of Genetics, Department of Medicine and Partners Center for Personalized Genetic Medicine, Brigham and Women’s Hospital and Harvard Medical School, Boston, Massachusetts, ^63^Department of Neurology, Massachusetts General Hospital/Harvard Medical School, Boston, Massachusetts, ^64^Department of Molecular Physiology and Biophysics, Vanderbilt University, Nashville, Tennessee, ^65^Vanderbilt Center for Human Genetics Research, Vanderbilt University, Nashville, Tennessee, ^66^Center for Applied Genomics, Children’s Hospital of Philadelphia, Philadelphia, Pennsylvania, ^67^Department of Pathology (Neuropathology), University of Pittsburgh, Pittsburgh, Pennsylvania, ^68^Institute of Neurology, University College London, Queen Square, London, ^69^Sanders-Brown Center on Aging, Department of Molecular and Biomedical Pharmacology, University of Kentucky, Lexington, Kentucky, ^70^Taub Institute on Alzheimer’s Disease and the Aging Brain, Department of Neurology, Columbia University, New York, New York, ^71^Department of Pathology, Duke University, Durham, North Carolina, ^72^Department of Genome Sciences, University of Washington, Seattle, Washington, ^73^Department of Medicine (Medical Genetics), University of Washington, Seattle, Washington, ^74^Sanders-Brown Center on Aging, Department Neurology, University of Kentucky, Lexington, Kentucky, ^75^Department of Pathology and Laboratory Medicine, University of California Davis, Sacramento, California, ^76^Department of Biostatistics, Boston University, Boston, Massachusetts, ^77^Department of Ophthalmology, Boston University, Boston, Massachusetts, ^78^University of Pittsburgh Alzheimer’s Disease Research Center, Pittsburgh, Pennsylvania, ^79^Department of Biology, Brigham Young University, Provo, Utah, ^80^Department of Neurology, Oregon Health & Science University, Portland, Oregon, ^81^Department of Neurology, Portland Veterans Affairs Medical Center, Portland, Oregon, ^82^Department of Pathology and Laboratory Medicine, University of California Irvine, Irvine, California, ^83^Department of Neurology, Boston University, Boston, Massachusetts, ^84^Department of Pathology, Boston University, Boston, Massachusetts, ^85^Department of Neuropsychology, University of California San Francisco, San Francisco, California, ^86^Department of Molecular & Medical Genetics, Oregon Health & Science University, Portland, Oregon, ^87^Department of Epidemiology, University of Washington, Seattle, Washington, ^88^Department of Neurobiology and Behavior, University of California Irvine, Irvine, California, ^89^Group Health Research Institute, Group Health, Seattle, Washington, ^90^Department of Pathology, University of Washington, Seattle, Washington, ^91^Department of Psychiatry and Behavioral Sciences, University of Washington, Seattle, Washington, ^92^Department of Pathology, University of Michigan, Ann Arbor, Michigan, ^93^Department of Psychiatry, Johns Hopkins University, Baltimore, Maryland, ^94^Department of Preventive Medicine, University of Southern California, Los Angeles, California, ^95^Department of Medicine - Pulmonary, New York University, New York, New York, ^96^Department of Neurology, University of Miami, Miami, Florida, ^97^Department of Pathology, University of California San Diego, La Jolla, California, ^98^School of Nursing Northwest Research Group on Aging, University of Washington, Seattle, Washington, ^99^Cognitive Neurology and Alzheimer’s Disease Center, Northwestern University, Chicago, Illinois, ^100^Department of Pathology, University of Southern California, Los Angeles, California, ^101^Department of Neurology, Washington University, St. Louis, Missouri, 102Department of Biostatistics, Mayo Clinic, Rochester, Minnesota, ^103^Department of Anatomic Pathology, Mayo Clinic, Rochester, Minnesota, ^104^Department of Laboratory Medicine and Pathology, Mayo Clinic, Rochester, Minnesota, ^105^Arizona Alzheimer’s Consortium, Phoenix, Arizona, ^106^Department of Psychiatry, University of Arizona, Phoenix, Arizona, ^107^Banner Alzheimer’s Institute, Phoenix, Arizona, ^108^Alzheimer’s Disease Center, New York University, New York, New York, ^109^Gertrude H. Sergievsky Center, Columbia University, New York, New York, ^110^Department of Neurology, Columbia University, New York, New York, ^111^Tanz Centre for Research in Neurodegenerative Disease, University of Toronto, Toronto, Ontario, ^112^Department of Neurology, University of Texas Southwestern, Dallas, Texas, ^113^Department of Radiology and Imaging Sciences, Indiana University, Indianapolis, Indiana, ^114^Department of Pathology (Neuropathology), Rush University Medical Center, Chicago, Illinois, ^115^Department of Psychiatry, University of Southern California, Los Angeles, California, ^116^Cambridge Institute for Medical Research and Department of Clinical Neurosciences, University of Cambridge, Cambridge, ^117^Department of Pathology, Johns Hopkins University, Baltimore, Maryland, ^118^Sanders-Brown Center on Aging, Department of Anatomy and Neurobiology, University of Kentucky, Lexington, Kentucky, ^119^Department of Pathology & Laboratory Medicine, University of California Los Angeles, Los Angeles, California, ^120^Taub Institute on Alzheimer’s Disease and the Aging Brain, Department of Pathology, Columbia University, New York, New York, ^121^Department of Psychiatry & Behavioral Sciences, Duke University, Durham, North Carolina, ^122^Department of Pathology, Oregon Health & Science University, Portland, Oregon, ^123^Evelyn F. McKnight Brain Institute, Department of Neurology, Miller School of Medicine, University of Miami, Miami, Florida

